# Long-term outdoor air pollution and COVID-19 mortality in London: an individual-level analysis

**DOI:** 10.1101/2023.02.16.23286017

**Authors:** Loes Charlton, Chris Gale, Jasper Morgan, Myer Glickman, Sean Beevers, Anna L Hansell, Vahé Nafilyan

## Abstract

**Background:** The risk of COVID-19 severity and mortality differs markedly by age, socio-demographic characteristics and pre-existing health status. Various studies have suggested that higher air pollution exposures also increase the likelihood of dying from COVID-19.

Objectives: To assess the association between long-term outdoor air pollution (NO_2_, NOx, PM_10_ and PM_2.5_) concentrations and the risk of death involving COVID-19, using a large individual-level dataset.

**Methods:** We used comprehensive individual-level data from the Office for National Statistics’ Public Health Data Asset for September 2020 to January 2022 and London Air Quality Network modelled air pollution concentrations available for 2016. Using Cox proportional hazard regression models, we adjusted for potential confounders including age, sex, vaccination status, dominant virus variants, geographical factors (such as population density), ethnicity, area and household-level deprivation, and health comorbidities.

**Results:** There were 737,356 confirmed COVID-19 cases including 9,315 COVID-related deaths. When only adjusting for age, sex, and vaccination status, there was an increased risk of dying from COVID-19 with increased exposure to all air pollutants studied (NO_2_: HR 1.07 [95% confidence interval: 1.04-1.12] per 10 μg/m^3^; NOx: 1.05[1.02-1.09] per 20 μg/m^3^; PM_10_: 1.32[1.15-1.51] per 10 μg/m^3^; PM_2.5_: 1.29[1.12-1.49] per 5 μg/m^3^). However, after adjustment including ethnicity and socio-economic factors the HRs were close to unity (NO_2_: 0.98[0.90-1.06]; NOx: 0.99[0.94-1.04]; PM_10_: 0.95[0.74-1.22]; PM_2.5_: 0.90[0.67-1.20]). Additional adjustment for dominant variant or pre-existing health comorbidities did not alter the results.

**Conclusions:** Observed associations between long-term outdoor air pollution exposure and COVID-19 mortality in London are strongly confounded by geography, ethnicity and deprivation.

**Summary:** Using a large individual-level dataset, we found that a positive association between long-term outdoor air pollution and COVID-19 mortality in London did not persist after adjusting for confounders including population density, ethnicity and deprivation.

## Introduction

Coronavirus disease (COVID-19), caused by the infectious respiratory SARS-CoV-2 virus, affects individuals differently, ranging from asymptomatic to hospitalisation, intensive care, and death. Prominent risk factors for severe disease and death include older age and male sex [1], respiratory and other pre-existing health conditions [2, 3, 1], as well as socio-economic factors such as increased levels of deprivation and non-white ethnicity [1, 4]. Air pollution exposure is also a factor of interest, as it has been shown to be pro-inflammatory with impacts on the human immune system that may potentially affect infectivity and severity of respiratory disease [5].

Research into air pollution and COVID-19 outcomes generally found that greater levels of pollution were associated with increased risk of severe disease and COVID-19 death. This has been shown in both ecological studies (e.g., [6, 7, 8, 9], reviewed by [10, 11], up to June 2021) and individual-level studies [12, 13, 14, 15, 16, 17, 18, 19, 20, 21]. Individual-level studies can adjust well for factors such as age, sex and socio-economic status [22, 6]. Most individual-level studies investigated disease severity such as hospitalisation [13, 12, 15, 17, 14, 16, 21, 19], generally identifying increased risk levels of ≥13%, in particular for PM_2.5_ (particulate matter with a small aerodynamic diameter (≤2.5μm)), although two studies found a lower or no increased risk [21, 19]. To date, few individual-level studies have analysed the effect of long-term air pollution on mortality following COVID-19 disease. These studies again found positive associations [14, 18, 20, 21, 23, 24], albeit of a smaller magnitude (≤11% increased risk), except one study, where fully adjusting for confounders removed the positive association [19].

However, some of these studies included low numbers of cases [12, 14, 16, 19], some did not include a sample representative of the entire population [15, 19] or suffered from participation bias [17, 16, 19], some only investigated one pollutant [20], and some did not adjust for all of the known risk factors which include age, sex, ethnicity, deprivation and health comorbidities [13, 12, 14, 18, 21]. All of these risk factors have been shown to confound the positive relationship between air pollution and COVID-19 mortality [25, 6, 26, 27].

In this study, we examined the association between long-term ambient air pollution (NO_2_, NOx, PM_2.5_ and PM_10_) at place of residence and the risk of death involving COVID-19 in 737,356 confirmed COVID-19 cases in London between September 2020 and January 2022 using a unique dataset based on the 2011 Census data linked to National Health Service medical care records.

## Material and methods

### Study data

We used data from the Office for National Statistics (ONS) Public Health Data Asset (PHDA). The ONS PHDA is a linked dataset combining the 2011 Census, mortality records, the General Practice Extraction Service (GPES) data for pandemic planning and research, the Hospital Episode Statistics (HES), and national testing data from National Health Service (NHS) Test and Trace, for the clinically vulnerable and health care workers (termed ‘pillar 1’) and the general population (‘pillar 2’). To obtain NHS numbers for the 2011 Census, we linked the 2011 Census to the 2011-2013 NHS Patient Registers using deterministic and probabilistic matching, with an overall linkage rate of 94.6% [28]. All subsequent linkages were based on NHS number.

### Study population

The study population was restricted to usual residents living in London (2011 Census area code E12000007) at the beginning of the pandemic, alive as of 1 September 2020, registered with a general practitioner, enumerated at the 2011 Census, aged up to 100 years old in 2011 and not in a care home in 2019 (7,176,832 records, ~80% of the population of London, with higher coverage in older age groups.

Data were linked to population density data (people per square Km) per Lower layer Super Output Area (LSOA, 34,753 in total) recorded mid-2018 and to national testing data from the NHS Test and Trace programme [4], including all records of confirmed positive SARS-CoV-2 polymerase chain reaction (PCR) or lateral flow tests conducted in hospital or the community. For individuals with multiple tests, we took the first positive test record.

There were 1,398,976 confirmed positive COVID-19 cases in London between 1 September 2020 and 12 December 2021 (the latest date within our Test & Trace data) (https://coronavirus.data.gov.uk/), of which 756,363 (54.1%) were linked to the ONS PHDA. After applying our inclusion criteria, the final dataset entered into the analysis contained 737,356 individual records, including 9,315 COVID-related deaths (~68.2% of all London COVID deaths up until 18 January 2022). The index date is the date of the first positive COVID test. The follow-up time was the earliest of: end of study date; death involving COVID-19; death from other cause.

### Outcome

The outcome was death involving COVID-19 (either in or out of hospital), defined as confirmed or suspected COVID-19 death as identified by ICD-10 codes U07.1 or U07.2 mentioned anywhere on the death certificate.

### Exposure

The 2016 annual average air pollution concentrations for NO_2_, NOx, PM_2.5_ and PM_10_ were obtained from the London Data Store (https://data.london.gov.uk/dataset/london-atmospheric-emissions-inventory--laei--2016, further details in Supplementary material). At the time of analysis this was the latest available data at high spatial resolution (20m) that had already been linked to ONS data. The spatial variability in air pollutants across London is relatively stable in recent years. Therefore, the 2016 data were taken as a reasonable proxy for recent long-term exposure. Hazard ratios (HRs) for NO_2_ are expressed per 10 μg/m^3^ increase, NOx per 20 μg/m^3^, PM_10_ per 10 μg/m^3^, and PM_2.5_ per 5 μg/m^3^ (following increments used by the European Study of Cohorts and Air Pollution Effects (ESCAPE) [29, 30]).

### Covariates

We included covariates known to be risk factors for COVID-19 [3, 2, 1] and likely to confound the relationship between air pollution exposure and risk of death [25, 6, 26, 1, 27]: age, sex, COVID-19 vaccination status, geographical factors, ethnicity, and socio-economic factors. In an additional analysis, we adjusted for non-respiratory pre-pandemic health status, and pre-existing respiratory morbidity, which potentially lie on the causal pathway between air pollution and risk of death.

Details, calculation, and data source of all covariates are detailed in Table 1.

**Table 1.** Model and covariate calculation, details and data sources.

| Model name | Covariates | Categories/Details | Source |
| --- | --- | --- | --- |
| **Age** | Age | In years at time of positive test; 3rd order polynomial | 2011 Census |
| **+ Sex** | Sex | Male, Female | 2011 Census |
| **+ Vaccination** | Vaccination 1 | At least 14 days < positive test date | NIMS* |
|  | Vaccination 2 | At least 14 days < positive test date | NIMS* |
| **+ Geo-graphical** | Population density | People per square Km, 2nd order spline cutoff at the 0.99 probability quantile | mid-2018 LSOA data |
|  | Rural/Urban | Major Conurbation, City and Town, Town and Fringe, Village, Hamlets & Isolated Dwellings | Primary care data |
|  | Local authority (LA) code | 45 areas, one of which combined 11 LAs with low variance; Entered as stratification variable | Primary care data |
| **+ Ethnicity** | Ethnicity | White, South Asian, Black, Other; Aggregated from 7 categories | 2011 Census |
| **+ Socio-economic\*\*** | Adjusted household deprivation | Not deprived, Deprived in 1 domain, Deprived in 2 domains, Deprived in all domains | 2011 Census |
|  | Adjusted IMD decile | 1 - most deprived to 10 - least deprived | English IMD 2019 |
| **+ Variant** | Dominant COVID-19 variant | Unknown (<01-12-2020), Alpha (01-12-2020 to 16-05-2021), Delta (17-05-2021 to study end); Entered as stratification variable, not included in further models | *** |
| **+ Non-respiratory health** | Disability (day to day activities) | Limited a lot, Limited a little, Not limited | 2011 Census |
|  | Learning disability | None, Learning disability, Down's syndrome | Primary care and hospital data |
|  | Binary health conditions | Diabetes (none or Type 2), Blood cancer, Atrial fibrillation, Congestive cardiac failure (CCF), Coronary heart disease (CHD), Congenital heart disease, Peripheral vascular disease (PVD), Chronic kidney disease (CKD, none or any stage), Stroke, Dementia, Rheumatoid arthritis or systemic lupus erythematosus, Liver cirrhosis | Primary care and hospital data |
|  | Body-mass index (BMI) | Underweight, Ideal, Overweight, Obese, Unknown | Primary care and hospital data |
| **+ All health** | Binary health conditions | Respiratory cancer, Asthma, Chronic obstructive pulmonary disease (COPD), Pulmonary hypertension | Primary care and hospital data |

\*National Immunisation Management Service (NIMS), \*\*This is our primary model, \*\*\*E. Bubb, "Coronavirus (COVID-19) Infection Survey Technical Article: Impact of vaccination on testing positive in the UK: October 2021," 2021.

Both household-level deprivation (2011 Census) and area-level deprivation (English Index for Multiple Deprivation 2019 (IMD)) were adjusted and rescaled to exclude the health-related components which can be mediating variables in the air pollution-COVID-19 mortality association (Supplementary material).

Pre-existing health conditions were derived from the primary care and hospital data, in a similar way to those included in the QCovid risk model [31]. For Body-mass index (BMI) we included a category for missing data as 362,298 (47.9%) individuals had no valid value. Dominant COVID-19 variants were defined following [32].

### Statistical analyses

We estimated the association between air pollution measures and the risk of COVID-related death using Cox proportional hazard models, adjusted for potential confounding factors such as age, sex, COVID-19 vaccination, geographical, socio-economic, and dominant variant (Table 1). To assess the role of each group of confounding factors, we iteratively adjusted for each of these factors. Our primary model was adjusted for all confounding factors but not pre-existing health conditions (air pollution, age, sex, first vaccination, second vaccination, population density, rural-urban, ethnicity, household deprivation, area-level deprivation (IMD), stratified by local authority).

We also presented the results from a model adjusted for pre-existing health conditions. Whilst these could mediate the relationship between air pollution and the risk of COVID-19 death, they may also be a confounding factor, since chronic conditions may lead to poverty and therefore influence exposure to air pollution.

Because exposures to the four pollutants are highly correlated, we fitted the series of models separately for each pollutant.

For each pollutant, we additionally ran the primary models with an interaction term each, separately, for age, ethnicity and deprivation (i.e., primary model covariants + covariant * pollutant). For each interaction model, we applied a log-likelihood ratio test (LLRT) to assess whether adding the interaction term improved the model. For variables with significant LLRT statistics, we ran sensitivity analyses, repeating the primary models for each variable’s subgroups (for age, binarized as below vs. from 65 years old).

Air pollution concentrations were linked to postcode of residence as at 2011 Census. As these may reflect outdated locations, we repeated all primary models using postcodes derived from the 2019 primary care patient register where available (35% differed from Census records).

The Cox Proportional Hazard models were performed using the Lifelines package for Python [33]).

## Results

### Characteristics of the study population

The analytical sample consisted of 737,356 individuals with a positive COVID test. They were aged 10-110 years (mean(SD): 39.4(18.7)), 54.2% were female. The sample included 9,315 COVID-related deaths (1.3%) (Table 2).

**Table 2.**
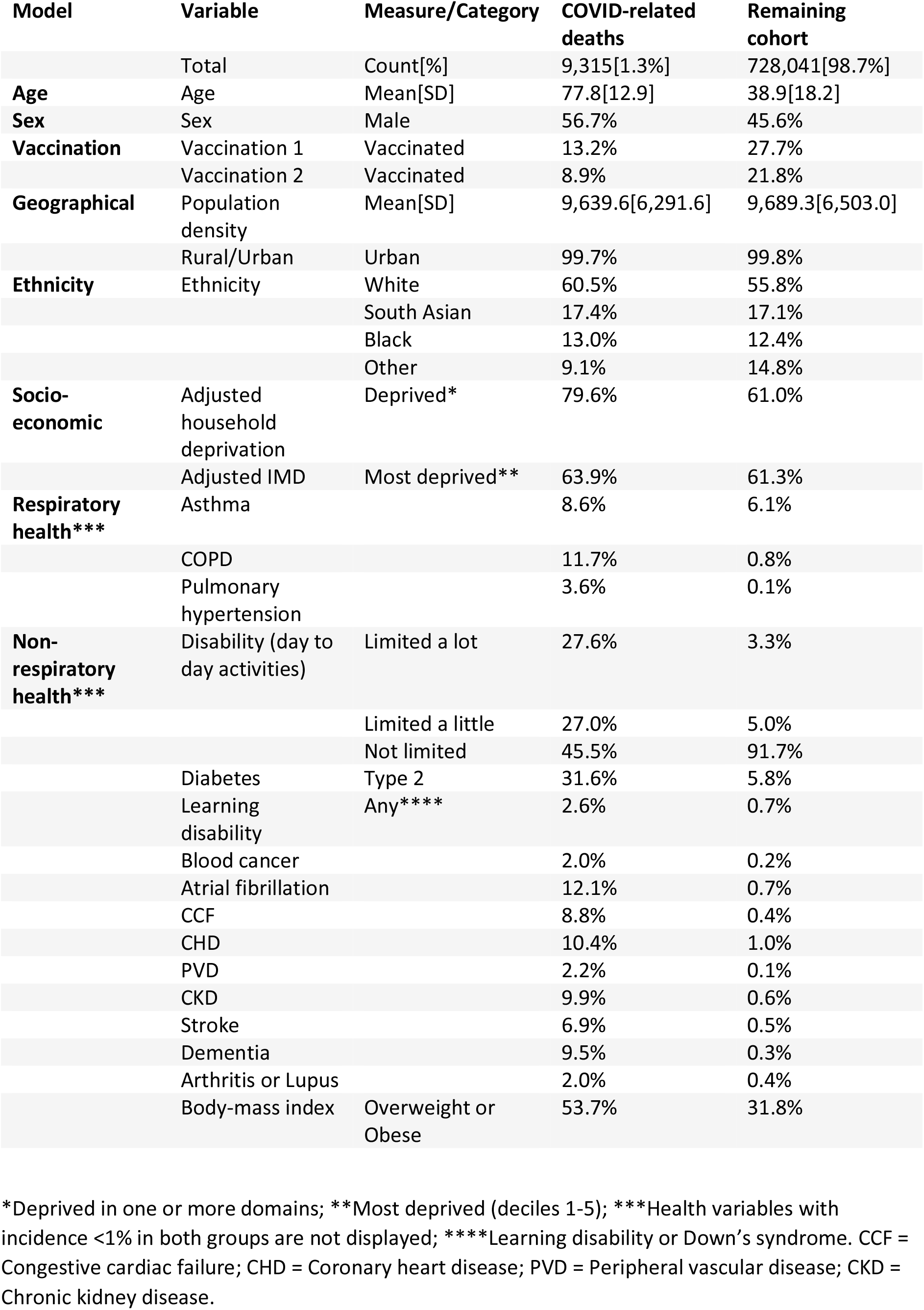
Study cohort and model details.

| Model | Variable | Measure/Category | COVID-related deaths | Remaining cohort |
| --- | --- | --- | --- | --- |
|  | Total | Count[%] | 9,315[1.3%] | 728,041[98.7%] |
| **Age** | Age | Mean[SD] | 77.8[12.9] | 38.9[18.2] |
| **Sex** | Sex | Male | 56.7% | 45.6% |
| **Vaccination** | Vaccination 1 | Vaccinated | 13.2% | 27.7% |
|  | Vaccination 2 | Vaccinated | 8.9% | 21.8% |
| **Geographical** | Population density | Mean[SD] | 9,639.6[6,291.6] | 9,689.3[6,503.0] |
|  | Rural/Urban | Urban | 99.7% | 99.8% |
| **Ethnicity** | Ethnicity | White | 60.5% | 55.8% |
|  |  | South Asian | 17.4% | 17.1% |
|  |  | Black | 13.0% | 12.4% |
|  |  | Other | 9.1% | 14.8% |
| **Socio-economic** | Adjusted household deprivation | Deprived* | 79.6% | 61.0% |
|  | Adjusted IMD | Most deprived** | 63.9% | 61.3% |
| **Respiratory health\*\*\*** | Asthma |  | 8.6% | 6.1% |
|  | COPD |  | 11.7% | 0.8% |
|  | Pulmonary hypertension |  | 3.6% | 0.1% |
| **Non-respiratory health\*\*\*** | Disability (day to day activities) | Limited a lot | 27.6% | 3.3% |
|  |  | Limited a little | 27.0% | 5.0% |
|  |  | Not limited | 45.5% | 91.7% |
|  | Diabetes | Type 2 | 31.6% | 5.8% |
|  | Learning disability | Any**** | 2.6% | 0.7% |
|  | Blood cancer |  | 2.0% | 0.2% |
|  | Atrial fibrillation |  | 12.1% | 0.7% |
|  | CCF |  | 8.8% | 0.4% |
|  | CHD |  | 10.4% | 1.0% |
|  | PVD |  | 2.2% | 0.1% |
|  | CKD |  | 9.9% | 0.6% |
|  | Stroke |  | 6.9% | 0.5% |
|  | Dementia |  | 9.5% | 0.3% |
|  | Arthritis or Lupus |  | 2.0% | 0.4% |
|  | Body-mass index | Overweight or Obese | 53.7% | 31.8% |

\*Deprived in one or more domains; \*\*Most deprived (deciles 1-5); \*\*\*Health variables with incidence <1% in both groups are not displayed; \*\*\*\*Learning disability or Down's syndrome. CCF = Congestive cardiac failure; CHD = Coronary heart disease; PVD = Peripheral vascular disease; CKD = Chronic kidney disease.

Whole-cohort 2016 annual average air pollution levels, median(IQR), were 34.3(6.4) µg/m^3^ for NO_2_, 58.7(16.4) µg/m^3^ for NOx, 21.5(2.0) µg/m^3^ for PM_10_ and 13.0(0.9) µg/m^3^ for PM_2.5_. Descriptive analyses did not suggest differences in air pollution levels comparing those who died with the surviving cohort members (Table 3). Correlations between pollutants were high (Table S1).

**Table 3.** Annual average air pollution concentrations at place of residence in 2016 (median[IQR] in µg/m^3^) broken down by variables known to vary with air pollution (geography, ethnicity and deprivation).

|  | NO<sub>2</sub> |  | NO<sub>x</sub> |  |
| --- | --- | --- | --- | --- |
| Cohort | COVID-related deaths | Remaining cohort | COVID-related deaths | Remaining cohort |
| Category |  |  |  |  |
| Major Conurbation (Most urban) | 34.0[6.3] | 34.4[6.4] | 57.8[16.2] | 58.8[16.4] |
| Hamlets & Isolated Dwellings (Most rural) | 24.7[1.2] | 26.3[4.4] | 36.0[4.0] | 40.2[9.8] |
| White ethnicity | 32.8[6.2] | 33.6[6.8] | 54.8[15.6] | 57.0[17.0] |
| South Asian ethnicity | 34.9[4.9] | 34.8[5.2] | 60.2[13.6] | 59.8[14.0] |
| Black ethnicity | 36.1[5.5] | 35.5[6.0] | 63.4[14.2] | 61.6[15.6] |
| Other ethnicity | 35.0[5.9] | 34.7[5.9] | 60.2[15.6] | 59.8[15.4] |
| Deprived in all domains (household) | 36.2[5.8] | 36.2[6.7] | 63.8[15.6] | 63.4[17.4] |
| Not deprived (household) | 33.3[5.5] | 33.6[6.0] | 56.0[13.8] | 56.8[15.0] |
| Most deprived (IMD decile 1) | 36.4[6.3] | 36.1[6.4] | 63.6[16.8] | 63.2[16.8] |
| Least deprived (IMD decile 10) | 30.5[4.2] | 31.0[4.7] | 49.2[10.5] | 50.4[11.4] |
|  | PM<sub>10</sub> |  | PM<sub>2.5</sub> |  |
| Cohort | COVID-related deaths | Remaining cohort | COVID-related deaths | Remaining cohort |
| Category |  |  |  |  |
| Major Conurbation (Most urban) | 21.3[2.0] | 21.5[2.0] | 13.0[0.8] | 13.0[0.9] |
| Hamlets & Isolated Dwellings (Most rural) | 18.4[0.5] | 19.0[1.6] | 11.5[0.2] | 11.8[0.8] |
| White ethnicity | 21.0[2.0] | 21.2[2.1] | 12.8[0.9] | 12.9[1.0] |
| South Asian ethnicity | 21.7[1.7] | 21.6[1.8] | 13.0[0.7] | 13.0[0.8] |
| Black ethnicity | 22.1[1.7] | 21.9[1.8] | 13.3[0.8] | 13.2[0.8] |
| Other ethnicity | 21.6[1.9] | 21.6[1.9] | 13.1[0.8] | 13.0[0.8] |
| Deprived in all domains (household) | 22.0[1.7] | 22.1[2.2] | 13.2[0.8] | 13.2[1.0] |
| Not deprived (household) | 21.1[1.8] | 21.2[1.8] | 12.8[0.8] | 12.8[0.8] |
| Most deprived (IMD decile 1) | 22.1[1.8] | 22.1[1.8] | 13.2[0.8] | 13.2[0.8] |
| Least deprived (IMD decile 10) | 20.2[1.5] | 20.4[1.5] | 12.5[0.6] | 12.5[0.6] |

Unadjusted mean air pollutant levels were compared via ANOVAs, showing significant differences across geographies, deprivation levels and ethnicities (Table S2).

### Modelling results

When only adjusted for age, sex, and vaccination status, all four pollutants were associated with an increased risk of death involving COVID-19 (Figure 1, Table S3) – results were for NO_2_ (HR[95% CI]: 1.07[1.03-1.12] per 10 μg/m^3^ increase), NOx (1.05[1.02-1.09] per 20 μg/m^3^), PM_10_ (1.32[1.15-1.51] per 10 μg/m^3^) and PM_2.5_ (1.29[1.12-1.49] per 5 μg/m^3^).

**Figure 1.**
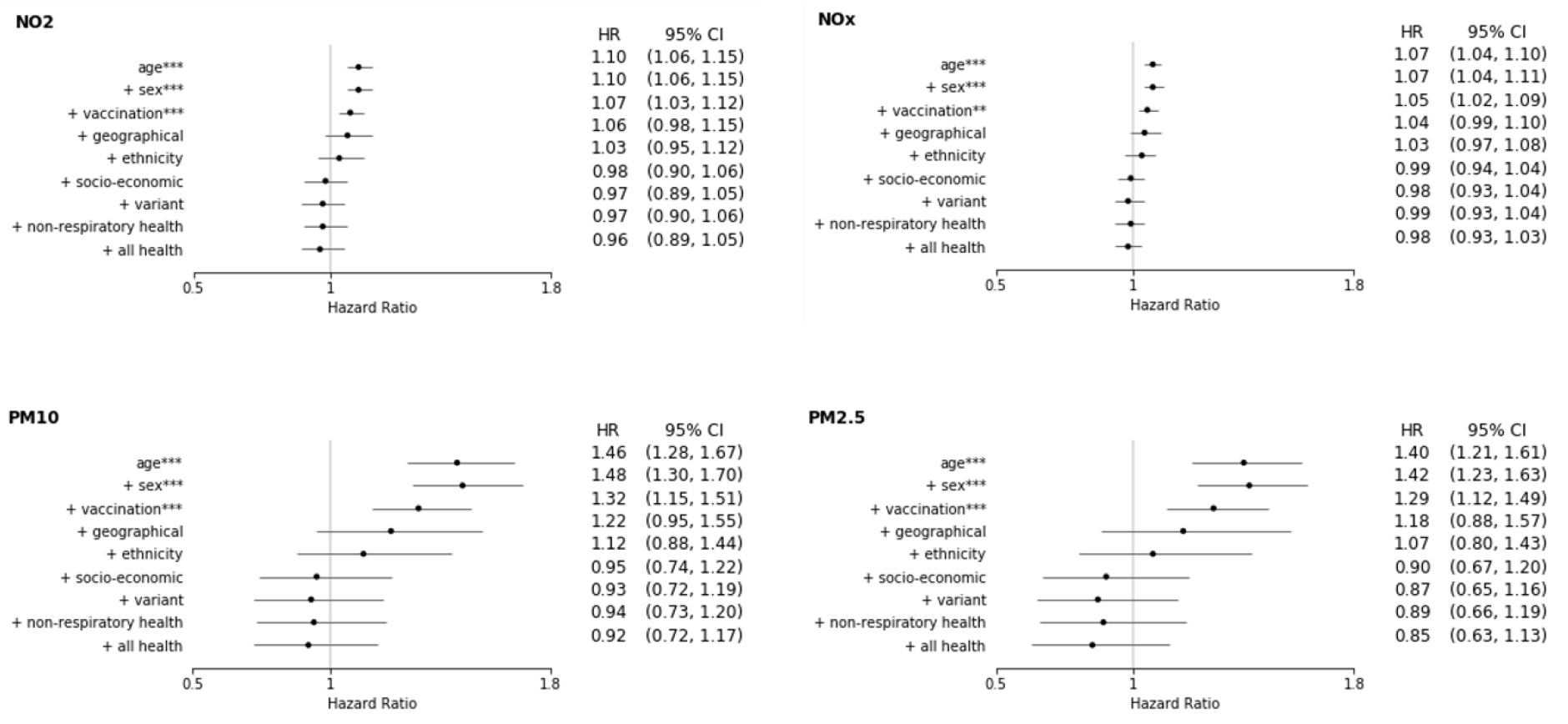
Hazard ratios (HRs) and 95% confidence intervals (CI) of each air pollution measure in the iterative models using 2011 residential address. The variables included in the models from top to bottom are subsequently additive, e.g., age, age + sex, age + sex + vaccination, etc. The exception is the ‘variant’ model. This model is equal to the previous model plus stratification by the dominant virus variant. The subsequent models including ‘health’ do not include this stratification. All models including geographical variables are stratified by local authority. *p<0.05, **p<0.01, ***p<0.001.

Results lost statistical significance and were slightly reduced in effect size when also adjusting the model for geographical factors (NO_2_1.06[0.98-1.15]; NOx 1.04[0.99-1.10]; PM_10_1.22[0.95-1.55]; PM_2.5_1.18[0.88-1.57]). After further adjusting for ethnicity and socio-economic factors, the HRs of all four pollutants were close to unity. Results remained null after additional adjustments for (i) dominant variant at the time of positive test, stratifying over the three variants; (ii) pre-pandemic health status of non-respiratory comorbidities; and (iii) respiratory comorbidities (Figure 1, Table S3).

We found significant interactions of air pollution with age for all pollutants, and ethnicity for all pollutants except NOx (Table S4). The only other significant interaction was household deprivation for PM_10_. We therefore conducted stratified analyses for age (below or from 65 years) and ethnicity (using main London ethnic groups of White, South Asian, Black, Other). While no HRs in stratified analyses were statistically significant, most were close to unity except for South Asian ethnicity where all air pollutants had positive associations with COVID-19 mortality, with the association for PM_2.5_ being HR[95%CI]: 1.40 [0.66-2.95] (Table S5).

Repeating the primary models using post-codes derived from the primary care patient register yielded highly similar main results (Figure S1).

## Discussion

### Main findings

In this analysis of almost three quarters of a million individuals in London who tested positive for COVID-19, we found that a positive association between long-term exposure to four air pollution measures (NO_2_, NOx, PM_10_ and PM_2.5_) and the risk of death involving COVID-19 became null after adjustment for geographical, ethnicity and socio-economic differences.

### Comparison with other studies

Most of the published work on this topic to date, based on both ecological [6, 7, 8, 9] and individual-level studies [13, 12, 15, 17, 16, 18, 14, 20, 21, 19] found that increased air pollution concentrations were associated with worse COVID-19 outcomes. However, there were some indications that ethnicity and deprivation [15, 19], as well as comorbidities, can greatly alter the air pollution-COVID-19 disease relationship [12, 17, 16].

Most published studies to date assessed outcomes such as severe symptoms or hospitalisation [13, 12, 15, 17, 16]. Only a small number of studies have investigated long-term air pollution and COVID-19 mortality: in Mexico city [18], California [20, 23, 24], New York city (NYC) [14], Canada [21], and England [19]. Their results were mixed.

Two of these studies did not adjust for ethnicity [18, 21]. The study from Mexico [18] found a 7.4% increased risk of dying per 1 μg/m^3^ increase in PM_2.5_, with the risk increasing with age. The study from Canada [21] only found an association between COVID-19 mortality and O_3_ (18% increased risk per interquartile range increase of 5.14 ppb).

The other studies did adjust for ethnicity. The study from NYC [14] on a small (N=6,542) but ethnically diverse hospitalised cohort found a positive COVID-mortality relationship for PM_2.5_ (11% increase per 1μg/m^3^ in adjusted models), though not for NO_2_ or black carbon. There was a trend for modification by ethnicity, but in stratified analyses, associations were only statistically significant for Hispanic, not for White, Black, or Asian ethnicities, despite reasonably similar air pollution concentration distributions. A large-sample study (N=3.1m) from California [20] identified a 3.8% increased risk per 1 μg/m^3^ increase in PM_2.5_ for their fully-adjusted model (including ethnicity and comorbidities). The effect was modified by both ethnicity and deprivation: there was a much greater risk for the Hispanic population and for those who were most deprived, though both groups lived in areas with greatest long-term PM_2.5_ exposure. Two other studies from Southern California found increased COVID-mortality risks for 1-year average non-freeway near-roadway air pollution (NRAP) (10% per 1SD) [23] and PM_2.5_ (14% per 1SD) [24] that did not interact with ethnicity. The study from England [19], covering a mainly White sample (average age 68), found that for PM_2.5_ and NO_2_ positive associations with COVID-mortality remained after adjusting for ethnicity, but disappeared once corrected for additional covariates such as deprivation.

There are differences between the design of our study and other mortality studies. Our study covered well over a year of the pandemic, starting after the first wave and including post-vaccine availability, whereas several previous individual-level studies covered mainly the first wave [18, 14, 20]. It is possible that the original Wuhan variant, pre-vaccination, induced more severe illness that was resulted in higher susceptibility to adverse effects of air pollution.

We used administrative test data to identify cases. However, universal availability of free testing in the UK and encouragement of case contacts to present for testing through the NHS mean that a large proportion of mild cases were detected. In our general-population study we had a COVID-19 fatality rate of 1.3%. This was higher than the voluntary-sample study in England (0.1%) [19], lower than the Mexico study (10-11%) [18], but equivalent to the general-population studies in California and Canada [20, 21]. In contrast, the New York study, which only considered patients admitted to hospital with COVID-19, had a fatality rate of 31% [14].

There were differences in confounder adjustments across studies. There is considerable evidence that the rate of COVID-19 mortality differs by ethnicity, in the UK and elsewhere [4, 1]. Both deprived and ethnic minority areas often experience higher air pollution exposures [34]. It is therefore imperative to consider both deprivation and ethnicity in COVID-19 analyses. However, the relationship between ethnicity and COVID-19 severity is complex and not only due to correlations with socio-economic status – there are documented genetic variants suggested to increase risks of COVID-19 severity that are more common in ethnic groups from South Asia [35], East Asia and the Americas [36].

### Strengths and limitations

The strengths of the current study are its large population size, a general population sample of those testing positive in London, and our comprehensive linkage of administrative microdata, including Census, hospital and primary care data, and accurate household-level deprivation data.

Furthermore, in analyses we distinguished between confounding and mediating variables, excluding the latter from our main model, and assessing their effect separately.

Results may not generalise to other geographical areas. The current study covered the area of Greater London, with people of different ages, ethnicity and socio-economic background generally relatively mixed across the city. Disentangling impacts of these factors can be difficult and London suffers to a degree from the ‘Manhattan effect’ (some wealthy areas in the city centre have highest levels of air pollution). Further, although the United Kingdom has a free National Healthcare system, COVID-19 test uptake is partially biased towards those with severe disease, and also likely influenced by occupation and socio-demographic factors [37]. However, we adjusted for both individual and area-level deprivation, and ethnicity was determined from Census self-report.

A further limitation is that some people may have relocated during the study period to a region with different air pollution levels, as we use data from the Census 2011. Repeating our analysis using 2011-2013 primary care registration data did not change the results (Fig. S1). However, there is still a possibility that a small number of patients may not have updated their registration data, resulting in misclassified location measurements.

We used the 2016 air pollution average as a proxy for long-term outdoor air pollution, which may not capture total exposure. At the time of analysis this was the most recently available measurement linked to our datasets. Air pollution levels may fluctuate over time, although the spatial distribution will remain relatively similar. Furthermore, other variables, such as occupation, place of work and type of daily transport likely contribute to total exposure. We also did not have data available to examine the contribution of short-term air pollution, which has been associated with increased COVID-19 mortality in other studies [38, 10, 11, 24, 23].

### Conclusion

Using linked administrative and health data to identify a large population-based pool of people testing positive for COVID-19, we found that initial observed associations between long-term outdoor air pollution exposure and the risk of dying from COVID-19 did not persist after adjusting for confounding factors, particularly deprivation and ethnicity. Impacts of air pollution on COVID-19 mortality have been observed in studies in other countries using different study designs and adjustment factors. Interpretations include that observed associations with air pollution are due to uncontrolled confounding; that close inter-relationships between confounders makes impacts of air pollution difficult to detect; and/or that air pollution effects are small in comparison with other risk factors such as deprivation and ethnicity. Lowering outdoor air pollution concentrations remains important with well-documented impacts on health and mortality, but our study suggests that to alleviate the mortality burden of COVID-19, the focus needs to be on improving the lives of those with lower socio-economic status and the circumstances in deprived areas [39].

## Data Availability

In accordance with NHS Digital's Information Governance requirements, the study data cannot be shared.

## Acknowledgements

ALH acknowledges support from the National Institute for Health Research (NIHR) Health Protection Research Unit (HPRU) in Environmental Exposures and Health at the University of Leicester development award, a partnership between the UK Health Security Agency, the Health and Safety Executive and the University of Leicester. The views expressed are those of the author(s) and not necessarily those of the NHS, the NIHR, the Department of Health and Social Care, the Health and Safety Executive or the UK Health Security Agency.

