## Supplementary Material for "Long-term outdoor air pollution and COVID-19 mortality in London: an individual-level analysis"

#### Title

* Office for National Statistics, Health Analysis and Pandemic Insights, Newport, NP10 8XG, UK;

** Environmental Research Group, School of Public Health, Imperial College London & MRC Centre for Environment and Health, Imperial College London;

*** Centre for Environmental Health and Sustainability, University of Leicester & NIHR Health Protection Research Unit in Environmental Exposures and Health at the University of Leicester & NIHR Health Protection Research Unit in Environmental Exposures and Health at the University of Leicester.

[1] Joint senior author



#### Abstract

Background: The risk of COVID-19 severity and mortality differs markedly by age, socio-demographic characteristics and pre-existing health status. Various studies have suggested that higher air pollution exposures also increase the likelihood of dying from COVID-19.

Objectives: To assess the association between long-term outdoor air pollution ($NO_2$, NOx, $PM_{10}$ and $PM_{2.5}$) concentrations and the risk of death involving COVID-19, using a large individual-level dataset.

Methods: We used comprehensive individual-level data from the Office for National Statistics' Public Health Data Asset for September 2020 to January 2022 and London Air Quality Network modelled air pollution concentrations available for 2016. Using Cox proportional hazard regression models, we adjusted for potential confounders including age, sex, vaccination status, dominant virus variants, geographical factors (such as population density), ethnicity, area and household-level deprivation, and health comorbidities.

Results: There were 737,356 confirmed COVID-19 cases including 9,315 COVID-related deaths. When only adjusting for age, sex, and vaccination status, there was an increased risk of dying from COVID-19 with increased exposure to all air pollutants studied ($NO_2$: HR 1.07 [95% confidence interval: 1.04-1.12] per 10 $\mu g/m^3$; NOx: 1.05[1.02-1.09] per 20 $\mu g/m^3$; $PM_{10}$: 1.32[1.15-1.51] per 10 $\mu g/m^3$; $PM_{2.5}$: 1.29[1.12-1.49] per 5 $\mu g/m^3$). However, after adjustment including ethnicity and socio-economic factors the HRs were close to unity ($NO_2$: 0.98[0.90-1.06]; NOx: 0.99[0.94-1.04]; $PM_{10}$: 0.95[0.74-1.22]; $PM_{2.5}$: 0.90[0.67-1.20]). Additional adjustment for dominant variant or pre-existing health comorbidities did not alter the results.

Conclusions: Observed associations between long-term outdoor air pollution exposure and COVID-19 mortality in London are strongly confounded by geography, ethnicity and deprivation.

### Supplementary material

#### Additional Material and Methods

##### Exposure details

The London Atmospheric Emissions Inventory (LAEI) model outputs were created using a dispersion modelling system incorporating ADMS dispersion model v5.2 (CERC, 2016) and road source model v5 (CERC, 2020), measured hourly meteorological data, empirically derived NO-$NO_2$-$O_3$ and PM relationships and emissions from the LAEI. The LAEI model has been instrumental in air quality decision making in London for schemes such as Congestion Charging and more recently for the London Mayor's Ultra Low Emissions Zone. The LAEI model gives annual mean air quality predictions of NO-$NO_2$-$O_3$, $PM_{10}$ and $PM_{2.5}$ on a regular 20m x 20m grid. Data for this study was taken from the London Data Store (https://data.london.gov.uk/dataset/london-atmospheric-emissions-inventory--laei--2016).

Comparison with London measurements results in the following good model performance:

**Model performance table. Values of the spearman correlation coefficient (r), root mean square error (RMSE), normalised mean gross error (NMGE) and normalised mean bias (NMB) for observed vs. modelled annual average concentrations for the year 2016.**

| Year | Pollutant | Number of monitoring sites (n) | Normalised mean bias (NMB) | Normalised mean gross error (NMGE) | Spearman correlation coefficient (r) |
|---|---|---|---|---|---|
| 2016 | NO2 | 97 | -0.05 | 0.25 | 0.81 |
| 2016 | NOx | 97 | -0.01 | 0.17 | 0.80 |
| 2016 | PM10 | 22 | 0.13 | 0.15 | 0.80 |
| 2016 | PM2.5 | 79 | 0.17 | 0.21 | 0.63 |

##### Adjusted deprivation calculation

For household deprivation, we calculated how many of the remaining three deprivation sub-domains (education, housing and employment) a household contained (1=not deprived, 2=deprived in one domain, 3=deprived in 2 domains, 4=deprived in all domains). We calculated an adjusted IMD by combining the standardised scores of the remaining 6 domains adjusting the original weighting to achieve a total of 100% (the excluded Health, Deprivation and Disability domain was weighted 13.5%, so we multiplied the remaining domain's weights by 100/86.5) (McLellan et al., 2019). We then converted this adjusted IMD to deciles (1=most deprived, 10=least deprived).

Table S1. Correlation matrix across whole-cohort mean pollutant levels (Pearson's correlation coefficient).

| | $NO_2$ | NOx | $PM_{10}$ | $PM_{2.5}$ |
|---|---|---|---|---|
| **$NO_2$** | 1.00 | 0.99 | 0.97 | 0.97 |
| **NOx** | 0.99 | 1.00 | 0.95 | 0.95 |
| **$PM_{10}$** | 0.97 | 0.95 | 1.00 | 0.98 |
| **$PM_{25}$** | 0.97 | 0.95 | 0.98 | 1.00 |

Table S2. ANOVA results comparing variable category means across the whole cohort for each pollutant. All F test results are displayed with df: $F_{4,\ 737351}$, with all $p<0.001$.

| | **Geography (Rural/urban)** | **Adjusted household deprivation** | **Adjusted IMD deciles** | **Ethnicity** |
|---|---|---|---|---|
| **$NO_2$** | 2261.54 | 23071.05 | 65419.67 | 5487.59 |
| **NOx** | 1745.39 | 21121.99 | 56616.37 | 4556.06 |
| **$PM_{10}$** | 2596.34 | 24117.41 | 73883.59 | 5709.66 |
| **$PM_{2.5}$** | 2698.69 | 21612.59 | 62331.15 | 4825.11 |

Table S3. Hazard ratios (HRs) and 95% confidence intervals (CI) of each air pollution measure in the iterative models (equivalent to Figure 1). The variables included in the models from top to bottom are subsequently additive, e.g., age, age + sex, age + sex + vaccination, etc. The exception is the 'variant' model. This model is equal to the previous model plus stratification by the dominant virus variant. The subsequent models including 'health' do not include this stratification. All models including geographical variables are stratified by local authority.

| $NO_2$ | HR | 95% CI low | 95% CI high | p value |
|---|---|---|---|---|
| Age | 1.10 | 1.06 | 1.15 | 0.0000 |
| + sex | 1.10 | 1.06 | 1.15 | 0.0000 |
| + vaccination | 1.07 | 1.03 | 1.12 | 0.0009 |
| + geographical | 1.06 | 0.98 | 1.15 | 0.1537 |
| + ethnicity | 1.03 | 0.95 | 1.12 | 0.4228 |
| + socio-economic | 0.98 | 0.90 | 1.06 | 0.6023 |
| + variant | 0.97 | 0.89 | 1.05 | 0.4609 |
| + non-respiratory health | 0.97 | 0.90 | 1.06 | 0.5178 |
| + all health | 0.96 | 0.89 | 1.05 | 0.3815 |

| NOx | HR | 95% CI low | 95% CI high | p value |
|---|---|---|---|---|
| age | 1.07 | 1.04 | 1.10 | 0.0000 |
| + sex | 1.07 | 1.04 | 1.11 | 0.0000 |
| + vaccination | 1.05 | 1.02 | 1.09 | 0.0013 |
| + geographical | 1.04 | 0.99 | 1.10 | 0.1332 |
| + ethnicity | 1.03 | 0.97 | 1.08 | 0.3381 |
| + socio-economic | 0.99 | 0.94 | 1.04 | 0.7046 |
| + variant | 0.98 | 0.93 | 1.04 | 0.5691 |
| + non-respiratory health | 0.99 | 0.93 | 1.04 | 0.5940 |
| + all health | 0.98 | 0.93 | 1.03 | 0.4480 |

| PM10 | HR | 95% CI low | 95% CI high | p value |
|---|---|---|---|---|
| age | 1.46 | 1.28 | 1.67 | 0.0000 |
| + sex | 1.48 | 1.30 | 1.70 | 0.0000 |
| + vaccination | 1.32 | 1.15 | 1.51 | 0.0001 |
| + geographical | 1.22 | 0.95 | 1.55 | 0.1167 |
| + ethnicity | 1.12 | 0.88 | 1.44 | 0.3532 |
| + socio-economic | 0.95 | 0.74 | 1.22 | 0.6901 |
| + variant | 0.93 | 0.72 | 1.19 | 0.5397 |
| + non-respiratory health | 0.94 | 0.73 | 1.20 | 0.6150 |
| + all health | 0.92 | 0.72 | 1.17 | 0.4871 |

| PM2.5 | HR | 95% CI low | 95% CI high | p value |
|---|---|---|---|---|
| age | 1.40 | 1.21 | 1.61 | 0.0000 |
| + sex | 1.42 | 1.23 | 1.63 | 0.0000 |
| + vaccination | 1.29 | 1.12 | 1.49 | 0.0005 |
| + geographical | 1.18 | 0.88 | 1.57 | 0.2728 |
| + ethnicity | 1.07 | 0.80 | 1.43 | 0.6685 |
| + socio-economic | 0.90 | 0.67 | 1.20 | 0.4681 |
| + variant | 0.87 | 0.65 | 1.16 | 0.3451 |
| + non-respiratory health | 0.89 | 0.66 | 1.19 | 0.4177 |

| | | | | |
|---|---|---|---|---|
| + all health | 0.85 | 0.63 | 1.13 | 0.2661 |

Table S4. Log-likelihood ratio tests for added interaction terms. The full model is our primary model plus the interaction term: air pollution + age + sex + vaccination + geographical + ethnicity + deprivation + air pollution * interaction variable. The reduced model is our primary model: air pollution + age + sex + vaccination + geographical + ethnicity + deprivation. LL = log likelihood, LLRT = log-likelihood ratio test.

| **Pollutant** | **Interaction variable** | **LL full model** | **LL reduced model** | **LLRT statistic** | **LLRT p value** |
|---|---|---|---|---|---|
| $NO_2$ | Age | -72985.9 | -73004.0 | 36.2 | 0.0000 |
| NOx | Age | -72989.3 | -72999.0 | 19.5 | 0.0001 |
| $PM_{10}$ | Age | -72984.3 | -73004.0 | 39.6 | 0.0000 |
| $PM_{2.5}$ | Age | -72988.5 | -73003.9 | 30.8 | 0.0000 |
| $NO_2$ | Household deprivation | -73003.9 | -73004.0 | 0.1 | 0.9456 |
| NOx | Household deprivation | -73004.0 | -72999.0 | -9.9 | 1.0000 |
| $PM_{10}$ | Household deprivation | -72999.9 | -73004.0 | 8.4 | 0.0153 |
| $PM_{2.5}$ | Household deprivation | -73003.9 | -73003.9 | 0.0 | 0.9946 |
| $NO_2$ | Ethnicity | -73000.0 | -73004.0 | 8.0 | 0.0184 |
| NOx | Ethnicity | -73000.6 | -72999.0 | -3.0 | 1.0000 |
| $PM_{10}$ | Ethnicity | -72995.6 | -73004.0 | 16.8 | 0.0002 |
| $PM_{2.5}$ | Ethnicity | -72986.6 | -73003.9 | 34.6 | 0.0000 |
| $NO_2$ | IMD decile | -73002.8 | -73004.0 | 2.3 | 0.3118 |
| NOx | IMD decile | -73003.8 | -72999.0 | -9.6 | 1.0000 |
| $PM_{10}$ | IMD decile | -73004.0 | -73004.0 | 0.0 | 0.9880 |
| $PM_{2.5}$ | IMD decile | -73001.1 | -73003.9 | 5.6 | 0.0604 |

Table S5. Analysis results stratified by age and ethnicity. Results displayed of our primary Cox proportional hazard model (air pollution + age + sex + vaccination + geographical + ethnicity + deprivation, leaving out the relevant stratified covariate). HR = Hazard ratio, CI = confidence interval.

| Pollutant | Category | HR (95% CI) | p value | Group size |
|---|---|---|---|---|
| $NO_2$ | Age < 65 years | 1.01 (0.83-1.22) | 0.9350 | 668,149 |
| NOx | Age < 65 years | 1.01 (0.90-1.14) | 0.8761 | |
| $PM_{10}$ | Age < 65 years | 0.97 (0.54-1.75) | 0.9154 | |
| $PM_{2.5}$ | Age < 65 years | 0.88 (0.43-1.80) | 0.7331 | |
| $NO_2$ | Aged 65+ years | 0.97 (0.89-1.06) | 0.5115 | 69,207 |
| NOx | Aged 65+ years | 0.98 (0.92-1.04) | 0.5126 | |
| $PM_{10}$ | Aged 65+ years | 0.95 (0.72-1.24) | 0.6872 | |
| $PM_{2.5}$ | Aged 65+ years | 0.91 (0.66-1.25) | 0.5589 | |
| $NO_2$ | White ethnicity | 0.99 (0.89-1.10) | 0.8265 | 411,605 |
| NOx | White ethnicity | 1.00 (0.93-1.07) | 0.9087 | |
| $PM_{10}$ | White ethnicity | 0.95 (0.69-1.32) | 0.7599 | |
| $PM_{2.5}$ | White ethnicity | 0.89 (0.61-1.29) | 0.5320 | |
| $NO_2$ | South Asian ethnicity | 1.06 (0.87-1.29) | 0.5469 | 125,783 |
| NOx | South Asian ethnicity | 1.02 (0.90-1.16) | 0.7152 | |
| $PM_{10}$ | South Asian ethnicity | 1.34 (0.72-2.50) | 0.3577 | |
| $PM_{2.5}$ | South Asian ethnicity | 1.40 (0.66-2.95) | 0.3829 | |
| $NO_2$ | Black ethnicity | 1.01 (0.82-1.25) | 0.8958 | 91,279 |
| NOx | Black ethnicity | 1.00 (0.88-1.15) | 0.9749 | |
| $PM_{10}$ | Black ethnicity | 1.02 (0.54-1.96) | 0.9433 | |
| $PM_{2.5}$ | Black ethnicity | 1.03 (0.46-2.30) | 0.9392 | |
| $NO_2$ | Other ethnicity | 0.88 (0.68-1.13) | 0.3112 | 108,689 |
| NOx | Other ethnicity | 0.93 (0.79-1.09) | 0.3511 | |
| $PM_{10}$ | Other ethnicity | 0.79 (0.37-1.71) | 0.5490 | |
| $PM_{2.5}$ | Other ethnicity | 0.76 (0.30-1.90) | 0.5549 | |

Figure S1. Hazard ratios (HRs) and 95% confidence intervals (CI) of each air pollution measure in the iterative models, calculated using postcodes derived from 2011-2013 primary care data. The variables included in the models from top to bottom are subsequently additive, e.g., age, age + sex, age + sex + vaccination, etc. The exception is the 'variant' model. This model is equal to the previous model plus stratification by the dominant virus variant. The subsequent models including 'health' do not include this stratification. All models including geographical variables are stratified by local authority. *p<0.05, **p<0.01, ***p<0.001.

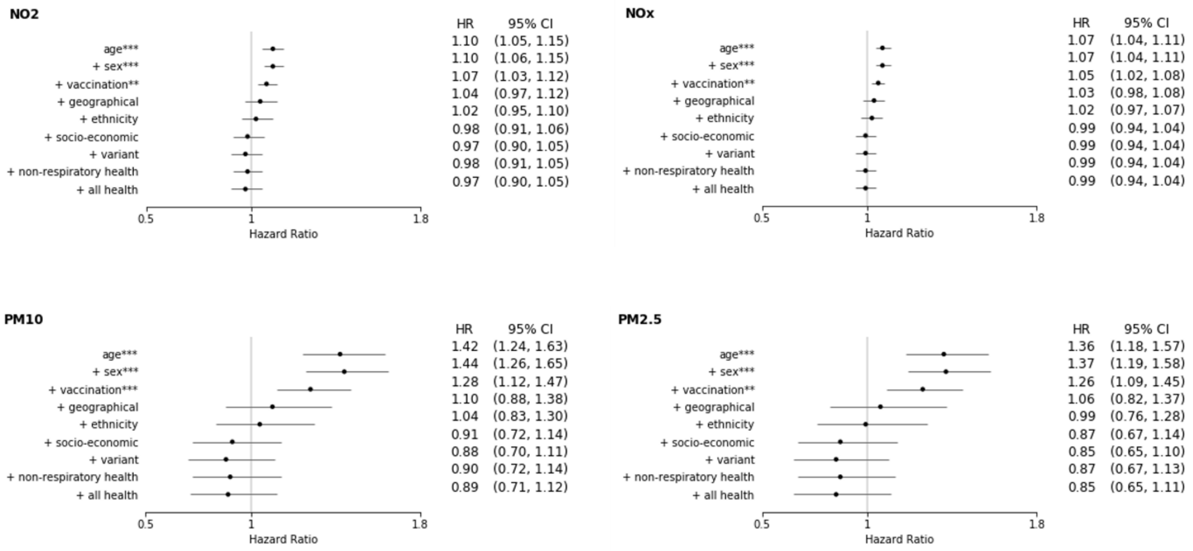
